# A whole-blood transcriptional signature associated with obstructive post-tuberculosis lung disease

**DOI:** 10.64898/2026.03.11.26348051

**Authors:** Renee E. Newby, Lucy Kijaro, Kimberly A. Dill-McFarland, Lilian N. Njagi, Wilfred B. Murithi, Zipporah Mwongera, Moeko Agata, Jodi Byers, Glenna J. Peterson, Kevin P. Fennelly, Videlis Nduba, David J. Horne, Jerry S. Zifodya, Thomas R. Hawn

## Abstract

**Background:** Post-tuberculosis (TB) lung disease (PTLD) affects approximately 50% of persons with pulmonary TB. We recently discovered whole blood transcriptional signatures associated with PTLD. We examined whether a minimum gene signature predicts PTLD as a clinically useful biomarker.

**Methods:** We prospectively enrolled 301 treatment naïve adults with newly diagnosed pulmonary TB (PTB) (cohort A). We collected whole blood at 0 and 6-month visits, isolated RNA, and measured a modified MTB Host Response (HR) signature (mHR) based on expression of DUSP3, GBP5, and TMBIM6. We recorded spirometry at 6 (n=216) and 12 months (n=210) after treatment initiation and examined the association of the mHR score with PTLD and Mtb aerosolization. We recruited household contacts of cohort A to compare mHR score with non-PTB participants (cohort B).

**Findings:** mHR was associated with TB (p=4.15e-66) when compared to HHCs, treatment response (p=1.07e-53), and characteristics including CD4 count (p=0.003), bacillary load (p=3.02e-05), lung cavities (p=1.59e-04), and lung quadrants involved (p=3.87e-06). The mHR score was not associated with Mtb aerosolization. In total, 105 (50%) participants had PTLD at 12 months including 61 with restriction, 26 with obstruction, and 18 with mixed obstruction and restriction. Baseline mHR was associated with obstructive PTLD at both 6 (p=0.003) and 12 months (p=0.012) in bivariate and multivariate analyses. The mHR score was not associated with restrictive lung disease.

**Interpretation:** Baseline mHR was associated with obstructive PTLD at 6 and 12 months and may have applications in targeting treatment and prognostication.

## Introduction

Tuberculosis (TB) is among the top 10 causes of death worldwide with an estimated 1.25 million deaths in 2023 and 10.8 million incident cases^1^. Whole blood transcriptional signatures are associated with pulmonary TB, treatment responses, and progression from latent to active pulmonary TB^2–8^. One signature, the Xpert MTB Host-Response assay (HR), initially identified by Sweeney et al.^6^, has been developed as a blood-based near-point of care assay run on the Cepheid Xpert platform. Xpert MTB HR predicts active TB and progression from latent to active TB, but has not been evaluated for predicting infectiousness or the development of post-TB lung disease (PTLD).

PTLD is a chronic respiratory abnormality attributable at least in part to previous tuberculosis,^9^ which affects roughly half of TB survivors ^10,11^. Predicting PTLD is important for prevention, prognostication, and stratification for new therapeutic interventions. There are currently no biomarkers or clinical algorithms for predicting the development of PTLD; however, we recently found whole blood transcriptional signatures based on RNA-Seq data that are associated with different PTLD endotypes^12^. This data suggests that a minimum gene transcriptional signature such as Xpert MTB HR could be developed as a PTLD biomarker.

People with pulmonary TB (PTB) who aerosolize Mtb are more likely to transmit TB to close contacts^11,12,13,14^. This association is stronger than other predictors of infectiousness, but its measurement is time and resource intensive. We previously discovered whole blood transcriptional profiles associated with Mtb aerosolization^15^. It is unknown if Xpert MTB HR or other minimal gene TB transcriptional signatures are associated with Mtb aerosolization.

To discover biomarkers and mechanisms of Mtb transmission and PTLD, we designed the TB aerobiology, immunology, and transmission (TBAIT) study in Nairobi, Kenya.^15,16^ The primary objectives of the current study were to identify associations between Xpert MTB HR with Mtb aerosolization, PTLD, levels of sputum and serum cytokines, and other clinical and laboratory features of PTB.

## Methods

### Study Design and Participants

We enrolled participants in a longitudinal cohort designed to assess TB aerobiology, immunology, and transmission (TBAIT) in Nairobi, Kenya^15^. Adults newly diagnosed with PTB by GeneXpert or liquid culture for Mtb were enrolled from outpatient clinics and through a door-to-door prevalence survey (index TB participants, Cohort A). All participants underwent sputum collection for GeneXpert (either MTB/RIF or Ultra) and two acid-fast bacilli (AFB) smears and culture (MGIT); participants were diagnosed with pulmonary TB based on a positive GeneXpert (Ultra >trace) or positive culture. All participants provided informed consent and completed a questionnaire that included data on symptoms, medical history, and socioeconomic factors. Household contacts (HHCs) of people with TB were enrolled if they resided in the same household for at least 60 days at the time of the index patient’s TB diagnosis (Cohort B). This study was approved by the Kenya Medical Research Institute (KEMRI)(Kenya Medical Research Institute) Scientific and Ethics Review Unit (SERU 048/3988), the Kenyatta National Hospital/University of Nairobi Ethics Review Committee (KNH-ERC/A/375) and the University of Washington Institutional Review Board (STUDY00009209). All participants provided written informed consent in either English or Kiswahili.

### Clinical and Laboratory Assessments

We administered a questionnaire to obtain clinical and demographic information, including age, sex, HIV status, alcohol use, smoking status, information on HHC sharing a bedroom with index, and anthropometric measures, including mid-upper arm circumference (MUAC), blood pressure (BP), weight, and height. Among index TB participants, HIV testing was performed using an opt-out approach, and CD4 lymphocyte count testing was performed where applicable. Chest radiographs (CXR) were obtained, and cavitation and quadrant involvement were assessed by a pulmonologist (DJH) and documented. We collected blood to measure white blood cell count (WBC), Hemoglobin-A1c, C-reactive protein (CRP), and a PAXgene tube for RNA analysis. Following diagnosis, participants were treated for 6 months with a four-drug regimen, (INH/rifampin/ethambutol/pyrazinamide) which was managed by the TB control program. Research follow-up visits at 6 and 12 months included symptom assessments, chest imaging, pulmonary function tests (PFTs), and additional blood samples for transcriptional analysis. HHCs underwent symptom screening (ongoing cough, hemoptysis, weight loss, chest pain, fevers, chills, or night sweats) and CXR to rule out active TB. HHCs with symptoms suggestive of TB and/or an abnormal CXR underwent sputum collection for GeneXpert. HHCs also completed a similar entry questionnaire, underwent laboratory tests including QuantiFERON-TB Plus (QFT-Plus, Qiagen Diagnostics; Hamburg, Germany), and provided blood samples for transcriptional analysis. HHCs were followed for one year to monitor for the development of TB.

### Cough Aerosol Cultures

Infectious aerosols were collected using a cough aerosol sampling system (CASS), as described previously^15^. Briefly, CASS consists of a six-stage Andersen Cascade Impactor (Thermo Fischer Scientific, IL) within a larger stainless-steel chamber attached to the mouthpiece tubing from the participant and with a vacuum pump, creating an airflow through the system^14,17^. Each of the six stages holds a Middlebrook plate with selective 7H10 or 7H11 solid agar media on which aerosolized particles settle based on size, with the smallest particles settling on the lowest plate. Participants were instructed to cough into a mouthpiece connected to a vacuum chamber that separates particles by size. CFUs are counted on each of 6 plates in the chamber. Plates were incubated at 37⍰°C and observed weekly for growth for a maximum of 8 weeks. Sputum samples were also collected and graded microscopically for the presence of AFB. GeneXpert MTB/RIF or GeneXpert Ultra, sputum cultures, and Ziehl-Neelsen (AFB) stains were performed on sputum samples. We additionally collected sputum for cytokine analyses.

### Pulmonary Function Tests

At the 6 and 12-month visit, participants returned for respiratory specific questionnaires and spirometry^18,19^. Spirometry was performed using the ultrasound-based ndd EasyOne^®^ Spirometer. Forced expiratory volume in one second (FEV1) and forced vital capacity (FVC) were measured according to ATS/ERS standards^20^. Each participant performed a minimum of three flow-volume loops before and after administration of a standard dose of an inhaled bronchodilator (salbutamol in four doses of 100µg by metered dose inhaler). The highest FEV1 and FVC from acceptable efforts for each participant were recorded. Global Lung Initiative (GLI) reference equations for “other” were used to determine predicted values^21^. In primary analyses, results were characterized as normal vs abnormal (indicating PTLD), using standard criteria to define restriction (FVC z-score <5^th^ percentile and FEV1/FVC > 5th percentile) and obstruction (FEV1/FVC<5th percentile; obstruction only and mixed phenotypes). FEV1 z-score, FVC z-score, and FEV1/FVC ratio were also modelled as continuous measures per ATS/ERS^22^.

### Whole blood transcriptional profile

Blood samples were collected in PAXgene tubes at enrollment (baseline) and end of TB treatment (month 6) and stored for later processing. PAXgene tubes were thawed at room temperature and RNA was isolated using PAXgene miRNA spin columns (Qiagen), followed by globin reduction using the GLOBINclear-Human kit (Thermo Fisher Scientific). RT-PCR and quantitative PCR were performed to obtain cycle thresholds of expression for DUSP3 and GBP5 as well as a housekeeping gene TMBIM6. This three-gene signature is modeled on the original Sweeney3 signature, which contained a different housekeeping gene (KLF2) and was derived for diagnosing TB with subsequent development as the Cepheid Host Response (HR) signature (also called Xpert MTB Host Response)^5–8^. Alternative housekeeping genes, including TMBIM6, have been developed with less heterogeneity among individuals^23,24^. We measured a modified Xpert MTB Host Response signature (mHR) with TMBIM6 as the housekeeping gene and calculated the score using the equation (CtDUSP3-CtTMBIM6 + CtGBP5-CtTMBIM6)/2. The score ranges from approximately -2 to 3 with a lower score denoting more inflammation and a stronger association with active TB.

### Data Analysis

All data were analyzed using R version 4.5. Statistical methods included Binomial and Gaussian generalized linear models to assess the relationships between variables. mHR was modeled as the independent variable with assessment of predictors including clinical variables, log-transformed CASS scores, PTLD phenotypes, and the risk of transmission to HHCs. To build an obstructive PTLD prediction tool, we used the Youden function of our area under the receiver operating characteristic curve (AUROC curve) analysis to select optimal cutoffs for continuous variables associated with obstructive PTLD at 6 and 12 months, converting them to binary variables. We included age given its biologically plausible association. We utilized a random forest algorithm to assess variable importance, implementing bootstrap subsampling with an ensemble of 500 trees, each using a random subset of 70% of the data, allowing for variability and reducing overfitting. We evaluated the importance of variables based on the Mean Decrease in Impurity (MDI), selecting those with importance scores greater than 1 for further analysis. This selection was applied to our cohort A, as illustrated in Supplemental Figure 1A & 1B.

## Results

### Association of mHR with Cohort A and Cohort B Characteristics

To examine whether mHR was associated with TB outcomes, we enrolled 301 participants with pulmonary TB (cohort A) with baseline (n=301) and end of treatment (month 6, n=149) whole blood RNA samples. The median age was 34 years with 20.9% women, 16% people living with HIV (PLWH), and 16.1% with diabetes (Previously diagnosed and/or confirmed HbA1c >6.5%) (Table 1). There was a significant difference between baseline mHR (median score = 0.034) and 6-month end of treatment mHR (median score = 1.24) (p 1.07e-53, Figure 1A). We also enrolled 217 participants who were HHCs (cohort B) of the pulmonary TB cases (cohort A). The median age for cohort B was 16 years with 62.7% women, 2.4% people living with HIV, and 1.1% with diabetes. The baseline mHR score was lower in those with pulmonary TB (median score=0.034) compared to HHCs (median score =1.34, p=4.15e-66, Figure 1B).

**Table 1.** Modified MTB Host Response Score Association with Pulmonary TB and Household Contacts Characteristics.

|  |  | Estimate | p-value |
| --- | --- | --- | --- |
| *A. Cohort A Pulmonary TB Cases (n = 301)* |  |  |  |
| Age (years) | 34 (26 - 41) | 0.004 | 0.144 |
| Women | 63 (20.9%) | 0.168 | 0.052 |
| BMI* (kg/m<sup>2</sup>) | 18.8 (17.3 - 20.7) | 0.041 | <0.001 |
| MUAC*, (cm) | 23 (21.3 - 24.9) | 0.053 | <0.001 |
| PLWH* (n<sub>total</sub>=205) | 33 (16.1%) | 0.028 | 0.795 |
| CD4 cell count, (n<sub>total</sub>=35)(cells/μL) | 225 (45.5 - 460) | 0.001 | 0.003 |
| HbA1c (mmol/mol) | 5.87 (5.5 - 6.21) | -0.017 | 0.407 |
| DM* | 48 (16.1) | -0.123 | 0.198 |
| Smear positive | 266 (88.4) | -0.093 | 0.396 |
| AFB* smear grade | 3 (2 - 4) | -0.051 | 0.056 |
| GeneXpert cycle threshold | 18 (16.4 - 21.8) | 0.029 | <0.001 |
| Number of involved quadrants on chest X-ray | 2 (1 - 3) | -0.146 | <0.001 |
| Cavitary disease | 207 (68.8) | -0.284 | <0.001 |
| *B. Cohort B Household Contacts (n = 217)* |  |  |  |
|  |  | Estimate | p-value |
| Age | 16.5 (6 - 30) | 0.003 | 0.160 |
| Women | 136 (62.7%) | 0.253 | <0.001 |
| BMI | 19.6 (16.6 - 25.3) | <0.001 | 0.861 |
| MUAC | 22.6 (16.8 - 27.2) | 0.005 | 0.415 |
| PLWH | 6 (2.76%) | -0.256 | 0.284 |
| DM | 3 (1.38%) | 0.089 | 0.760 |
| TB QuantiFERON | 194 (62.4%) | -0.066 | 0.372 |
| Cohort A CASS positivity | 52 (31.7) n = 164 | 0.029 | 0.734 |
| Cohort A contact PLWH | 10 (7.52%) (n=143) | -0.104 | 0.518 |
| Cohort A contact smear positivity | 183 (84.3%) | -0.013 | 0.570 |
| Cohort A smear grade | 3 (2 - 4) | -0.013 | 0.570 |
| Cohort A contact cavitary disease | 140 (64.5%) | -0.099 | 0.163 |
| Cohort A mHR score* | 0.009 (-0.449 - 0.325) | 0.057 | 0.314 |
| Cohort A CXR severity | 2 (1 - 2) | -0.284 | 0.360 |
| Cohort A GeneXpert CT value | 17.8 (16.4 - 21.6) | -0.003 | 0.697 |
| Cohort A CRP | 58.1 (34.6 - 95.8) | -0.001 | 0.406 |

Presented as median (interquartile range) and number (%), \*AFB=acid-fast bacilli; BMI=body mass index; CT=cycle threshold; CXR=chest X-ray; MUAC=mid upper arm circumference; DM=Diabetes Mellitus; PLWH=persons living with HIV; mHR=modified MTB Host Response score.

In Cohort A, low baseline mHR was associated with body mass index (lower BMI), MUAC, CD4 count, GeneXpert cycle threshold, and higher number of affected CXR quadrants, CRP, and presence of cavitary disease (Table 1, Figure 2A-D). In a multivariate regression, mHR remained associated with each of these variables except for the number of affected quadrants (Supplemental Table 1). There were no associations between mHR and other measured serum or sputum inflammatory markers (serum TNF-⍰ and IL-6 and sputum IL1B, CXCL8 and IL-6 (Supplemental Table 2). In Cohort B, mHR was associated with female sex (p < 0.001, Table 1). There was no association between cohort B baseline mHR scores with any features of cohort A including cohort A mHR scores (Table 1).

**Figure 1:**
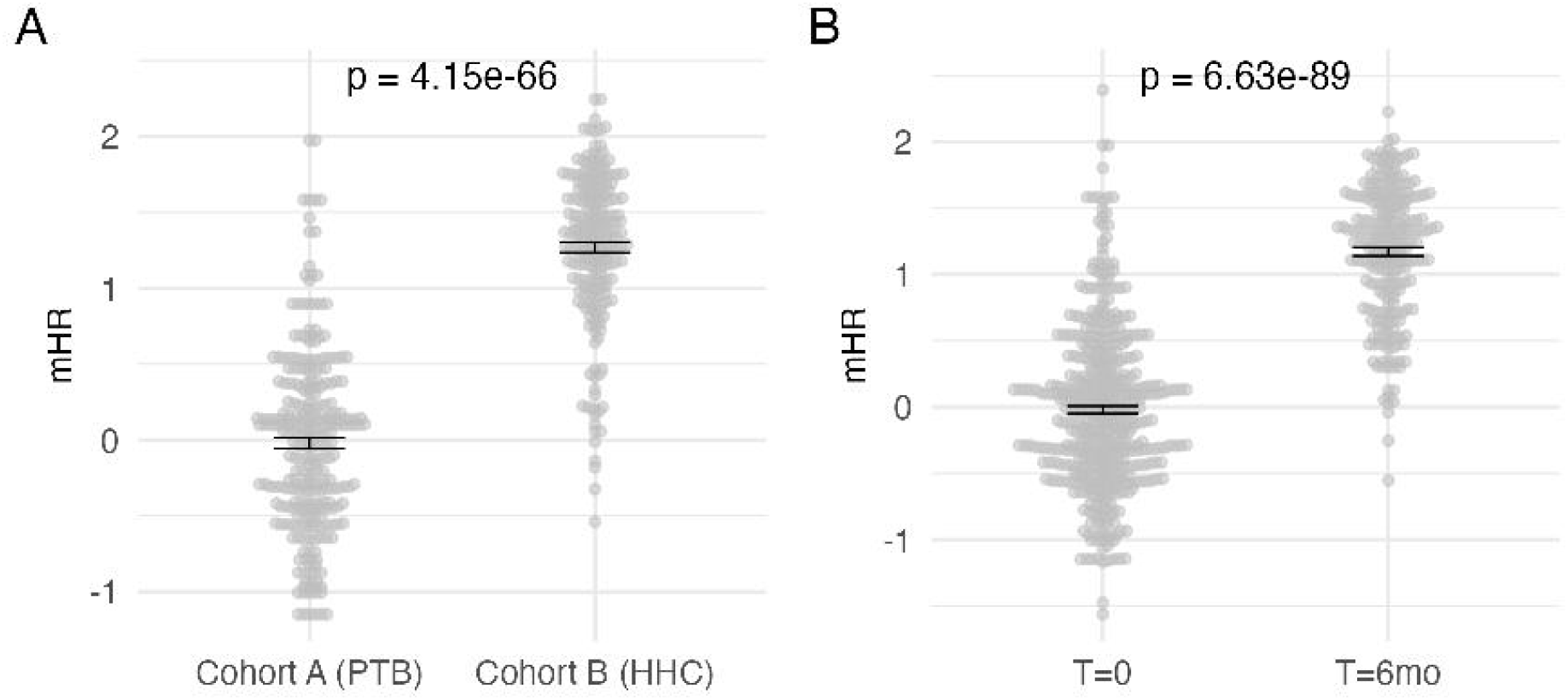
Association of mHR score with pulmonary TB and treatment. A. mHR scores in cohort A (PTB cases) and their household contacts (Cohort B). B. mHR scores in cohort A at the beginning (T=0) and end of a 6-month treatment course (T=6mo). p-values and correlation coefficients (R) calculated with a gaussian generalized linear model. Black bars indicate standard deviation around the mean.

**Figure 2:**
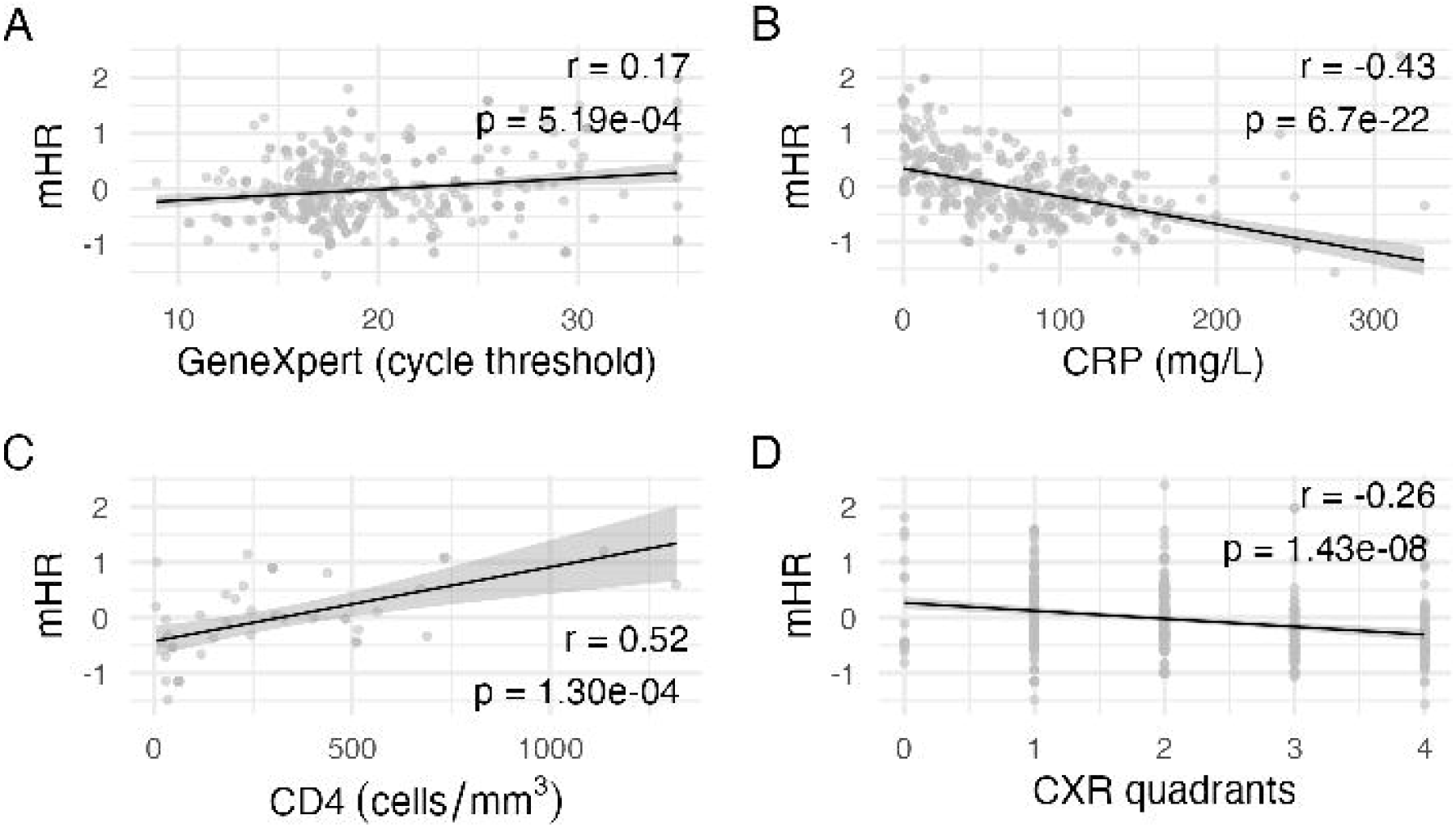
Associations between mHR and clinical and laboratory variables among pulmonary TB cases. Significantly associated variables are depicted with mHR scores in correlation plots including **A**. GeneXpert cycle threshold values (p=3.02e-05), **B**. CRP (p=1.05e-12 0.001), C. CD4 count (p = 0.003) and D. number of involved CXR quadrants (p=3.87e-06). p-values and correlation coefficients (R) calculated with a gaussian generalized linear model.

### mHR and Cough Aerosol Culture Status

We next examined whether mHR was associated with CAC positivity. Of the 301 cohort A subjects, we measured mHR scores on all CAC positive individuals (N=67) and 139 CAC negative participants (to enrich for CAC+ representation) (Supplemental Table 3). In individual bivariate regressions, low mHR and GeneXpert cycle threshold (CT) value, increased cavitary disease, and CRP were associated with CAC positivity (Supplemental Table 3). To avoid confounding, CRP was not carried forward in the multivariate analysis due to its collinearity with mHR. In a multivariate analysis with GeneXpert cycle threshold, cavitary disease and age, only GeneXpert remained significantly associated with CAC status (p =2.58e-04). There was no association between Cohort A CAC status and the mHR score of the associated HHC in cohort B.

### mHR and PTLD

To examine associations of mHR with PTLD, we measured pulmonary function tests at 6 and 12 months. Of the 301 cohort A participants, 216 had spirometry at 6 months and 210 had spirometry at 12 months. At 6 months, 106 (49%) had PTLD including 69 (65%) with restriction, 26 (25%) with obstruction, and 11 (10%) with mixed restriction and obstruction. At 12 months, 105 (50%) had PTLD including 61 (58%) with restriction, 26 (25%) with obstruction, and 18 (17%) with mixed obstruction and restriction. Baseline mHR was associated with obstructive PTLD at both 6 (p=0.003) and 12 months (p=0.012, Table 2, Figure 3A & 3B). In contrast, the 6-month mHR score was not associated with obstructive PTLD (Table 2). Interestingly, baseline mHR was not associated with the restrictive sub-phenotype or overall PTLD. Baseline mHR was the only significant association with 6-month obstruction (p = 0.003), while both mHR (p = 0.023) and number of affected CXR quadrants (p = 0.002) were associated with 12-month obstruction. In a multivariate analysis, higher baseline mHR remained significantly associated with obstruction at 12 months (p=0.023). Finally, the results of our 6-month PTLD obstruction prediction model including predictors(optimal AUC cutoffs); age(36.5), BMI(20.91), CXR quadrants(1.5), CRP(25.87) and mHR(0.83), yielded an AUROC of 0.781. Our 12-month prediction model included predictors; sex, age, mHR and CXR quadrants and yielded an AUROC of 0.820 (Figure 3C & 3D).

**Table 2.** Modified MTB Host Response Score Association with Obstructive PTLD at 6 and 12 months.

| *A. 6 month Pulmonary Function Test Assessment* |  |  |  |  |
| --- | --- | --- | --- | --- |
|  | Normal PFTs (n = 110) | Obstructive PFTs (n = 18) | OR | p-value |
| Age (years) | 32.5 (25 - 43) | 37.9 (21 - 74) | 1.018 | 0.356 |
| Sex (female) | 25 (22.7%) | 3 (16.7) | 0.680 | 0.566 |
| PLWH* | 10 (12.2%) | 1 (7.69) | 0.600 | 0.641 |
| DM* | 12 (10.9%) | 2 (11.1) | 1.021 | 0.980 |
| HgA1c (mmol/mol) | 5.7 (5.45 - 6.04) | 5.7 (4.76 - 7.13) | 0.809 | 0.476 |
| MUAC (cm) | 23.5 (22 - 25.5) | 22.8 (18 - 27) | 0.879 | 0.179 |
| BMI (kg/m<sup>2</sup>) | 19.8 (18.2 - 22.1) | 19 (15 - 24.2) | 0.825 | 0.071 |
| AFB Smear positive | 90 (81.8%) | 16 (88.9) | 1.778 | 0.444 |
| GeneXpert cycle threshold | 18.8 (17.2 - 24) | 20.7 (12 - 35) | 0.994 | 0.897 |
| Cavitary disease | 17 (15.5%) | 2 (11.1) | 0.684 | 0.633 |
| CRP (mg/L) | 49.8 (15.1 - 100) | 38.4 (0.2 - 131) | 0.989 | 0.075 |
| CXR severity | 2 (1 - 2) | 2.06 (1 - 3) | 1.685 | 0.074 |
| Baseline mHR Score | 0.037 (-0.334 - 0.581) | 0.699 (-0.160 - 1.97) | 5.232 | 0.003 |
| 6-month mHR | 1.33 (0.868 - 1.62) | 1.38 (1.28 - 1.55) | 1.268 | 0.692 |
| *B. 12 month Pulmonary Function Test Assessment* |  |  |  |  |
|  | Normal PFTs (n = 105) | Obstructive PFTs (n = 26) | OR | p-value |
| Age (years) | 31 (25 - 44) | 37.8 (21 - 74) | 1.017 | 0.335 |
| Sex (female) | 26 (24.8%) | 3 (11.5%) | 0.396 | 0.157 |
| PLWH | 10 (9.5%) | 2 (7.7%) | 0.695 | 0.655 |
| DM* | 10 (9.5%) | 0 (0) | <0.001 | 0.99 |
| HgA1c (mmol/mol) | 5.56 (5.3 - 5.98) | 5.65 (4.34 - 6.45) | 0.850 | 0.464 |
| MUAC (cm) | 23.5 (21.9 - 25.4) | 22.8 (18 - 27) | 0.867 | 0.118 |
| BMI (kg/m<sup>2</sup>) | 19.5 (17.8 - 21.3) | 19.1 (15 - 24.2) | 0.883 | 0.179 |
| AFB Smear positive | 89 (84.8%) | 24 (92.3) | 2.157 | 0.444 |
| GeneXpert cycle threshold | 19.4 (16.9 - 24.2) | 19 (8.9 - 35) | 0.940 | 0.165 |
| Cavitary disease | 20 (19.0%) | 4 (15.4) | 0.773 | 0.666 |
| CRP (mg/L) | 53.2 (15.1 - 91.8) | 51.5 (2.29 - 134) | 0.996 | 0.437 |
| CXR severity (quadrants) | 1 (1 - 2) | 2.27 (1 - 4) | 2.141 | 0.002 |
| Baseline mHR Score | -0.174 (-0.498 - 0.541) (61) | 0.403 (-0.934 - 1.97) | 2.496 | 0.023 |
| 6-month mHR | 1.33 (0.868 - 1.62) | 1.38 (1.28 - 1.55) (10) | 1.442 | 0.692 |

Presented as median (interquartile range) and number (%), \*AFB=acid-fast bacilli; BMI=body mass index; CT=cycle threshold; CXR=chest X-ray; MUAC=mid upper arm circumference; DM=Diabetes Mellitus; PLWH=persons living with HIV; mHR=modified MTB Host Response score.

**Figure 3:**
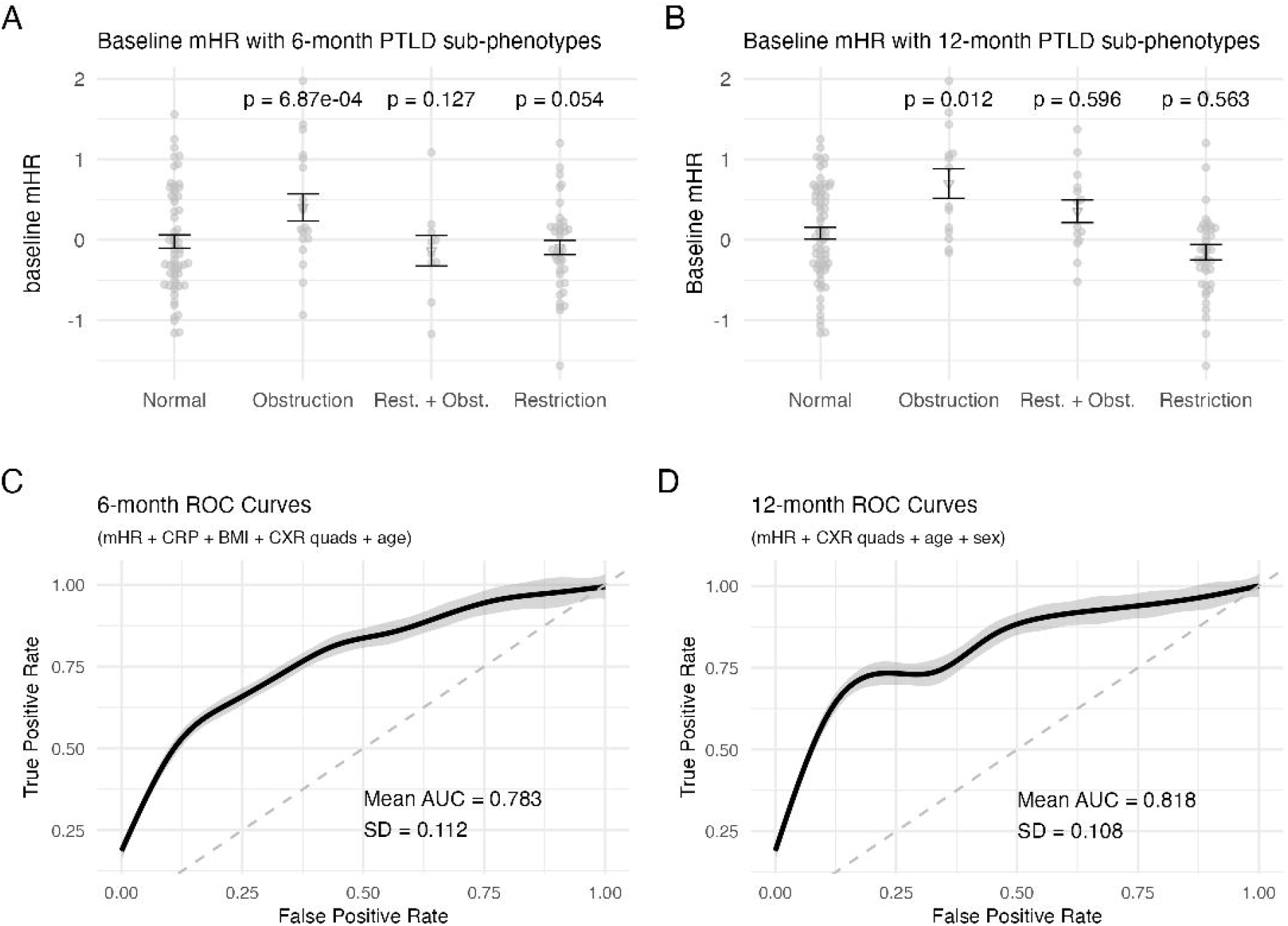
Association of mHR score with obstructive lung disease at 6 and 12 months. A and B. mHR score associations with PTLD phenotypes at 6 and 12 months compared to mHR of TB patients with normal lung function. Obstructive disease has a higher mHR score on average at both 6 (p=0.003) and 12 months (p=0.023), while mixed and restrictive phenotypes are not significantly different from normal at 6 or 12 months. p-values were calculated with a binomial generalized linear model. Black bars indicate standard deviation around the mean. The importance of discrete variables that may contribute to a prediction of obstructive disease at 6 or 12 months, respectively, was assessed by random forest variable importance scores. Variables with importance >1 were carried forward into random forest AUC analysis (Figure S1). **C and D**. The model predicted obstruction at 6 months with an AUC of 0.781 using mHR, CRP, BMI, age, and number of CXR quadrants. An AUC of 0.820 was achieved for predicting obstruction at 12 months using mHR, number of CXR quadrants, age and sex. The shaded area represents confidence intervals. ROC curves with mean area under the curve (AUC) and standard deviation (SD) were generated across cross-validated random forest models trained and tested on 100 random 80:20 data splits.

### Predictive model

Finally, we developed a predictive model with baseline characteristics associated with obstructive PTLD at 6 and 12 months using variables with p-values < 0.1, including mHR, sex, affected CXR quadrants, BMI and CRP. We also included age due to biological plausibility. We converted all continuous variables into binary variables with an optimal cutoff based on AUROC of each variable with the obstructive sub-phenotype. We additionally performed a random forest variable importance analysis of the above variables and included those with an importance > 1 (Supplemental Figure 1A & 1B). In the final model, age, BMI, CXR quadrants, CRP and mHR were included for the 6-month prediction with an overall model AUROC of 0.781. Sex, age, mHR and CXR quadrants were included for the 12-month prediction with an overall model AUROC of 0.820 (Figure 3C & 3D). Together, these data suggest that mHR is associated with obstructive, but not restrictive PTLD.

## Discussion

Modified Xpert MTB Host Response transcriptional signature at the time of TB diagnosis is a potential biomarker for obstructive PTLD assessed at 6 months following treatment completion. We also found associations of mHR with CD4 count, bacillary load, and the degree and nature of lung involvement. To our knowledge, the association with CD4 count and extent of lung involvement has not been reported previously. In contrast, we did not find associations of mHR with Mtb aerosolization in multivariate analyses.

PTLD includes obstructive and restrictive patterns representing distinct clinical phenotypes with largely unknown pathogenesis mechanisms^25^. The mHR signature contains GBP5 (Guanylate-Binding Protein 5), which is an interferon-inducible gene involved in antimicrobial defense and inflammation, and as such, may reflect activation of type 1 interferon responses^26,27^. Using whole blood RNASeq profiling of a smaller subset of the same TBAIT cohort, we recently found an association of an IFNα and IFNγ response signature at month 6 with PTLD^16^. We also found that restrictive PTLD was marked by early pro-fibrotic inflammation at the time of TB diagnosis (IL-6/JAK/STAT3 and TNF-αsignaling), while obstructive PTLD was not associated with any baseline differentially expressed genes (DEGs)^16^. Together, these mHR and RNA-Seq data suggest distinct inflammatory pathways associated with obstructive and restrictive PTLD with the possibility of persistent inflammation modulating obstructive PTLD. The finding that baseline mHR is associated with obstructive PTLD highlights a potential for early identification and targeted interventions for this specific PTLD phenotype. This work builds upon growing evidence of PTLD’s heterogeneity, where there are various phenotypes/clinical patterns^25^. PTLD is now further endotyped using gene expression data at TB diagnosis and after treatment completion^16^.

The etiology of smoking-related COPD has been examined with discovery of associated genetic, transcriptomic, and proteomic signatures^28–30^. One study found decreased expression of GBP5 in transcriptional profiles of subjects with COPD due to smoking^31,32^. Lower GBP5 expression was also observed in smokers with and with without COPD in a separate study^32,33^. These studies highlight possible overlapping inflammatory pathways associated with smoking-related COPD and obstructive PTLD. Further comparative studies of smoking-related COPD subjects with obstructive PTLD will be needed to uncover unique and overlapping inflammatory pathways.

mHR has been studied in several clinical contexts and various prior findings were confirmed in our study. Among them, we confirmed the difference in people with and without pulmonary TB (p < 0.001)^2,5–8,34–36^, pre and post-treatment response^7^, and associations with cavitary disease^5,34^, CRP^5,37^, MUAC/BMI^5^, and bacillary burden^5,34,38^. To our knowledge, previous studies have not reported the association of mHR with CD4 count, extent of disease as shown by the number of involved chest X-ray quadrants, or obstructive PTLD. While prior studies have included persons living with HIV, CD4 counts have been >200,^5^ which may obscure a potential relationship. Our minimum CD4 count was 29 with a median of 225, and those with low CD4 counts were more likely to have a lower mHR score. Interestingly, a key component of mHR is GBP5, which may play a role in viral and bacterial defense, and may have dual purposes in people living with HIV who are not on antiretroviral therapy^39^.

Our study has several limitations. First, our study was conducted in a single region of Kenya, which may limit the generalizability of our findings to other populations. However, by focusing on a population with a high burden of TB, we were able to power our study, providing insights into the relationship between mHR and TB disease. Second, the spirometry data were collected at time points relatively close to the period of TB treatment, a time with many dynamic lung changes. Although we anticipate further lung remodeling over time, the end of treatment and 6-month post treatment findings were similar. We are collecting 18- and 24-month pulmonary function tests and plan to analyze this data in future PTLD studies. Third, our sample size was modest for PTLD outcomes. Despite this, the sample was sufficiently powered to detect statistically significant associations. Finally, the mHR signature was not specifically developed as a biomarker for PTLD. One of our next steps will be to derive a de novo minimum gene signature for PTLD from our RNA sequencing data and then test it with qPCR in the TBAIT cohort as a potential refined PTLD biomarker. Future research is needed to validate these findings in larger, independent cohorts and to elucidate the specific mechanisms underlying mHR’s association with the development of obstructive PTLD. Future studies could also explore whether interventions targeting the pathways associated with mHR can reduce the incidence or severity of obstructive PTLD

## Data Availability

All data produced in the present study are available upon reasonable request to the authors

## Acknowledgments

We would like to thank the individual study participants and their families. We thank Dr Joy Githua, Dr Jacqueline Mirera, Robi Chacha, Lenis Njagi, Geoffrey Onchiri, Ruth Munyasya, Caroline Epiche, Isaac Kibet, Stella Nthambi, Kevin Munge, Japherson Mecha, Patrick Isinidu, Joash Omolo, Hastings Koech, Inviolata Sakwa and all other KEMRI CRDR Nairobi staff for their support in data collection. We thank Simon C. Mendelsohn and Thomas J. Scriba for advice on transcriptomic analyses.

## Citations

1. Global Tuberculosis Report 2024. (World Health Organization, Geneva, 2024).

2. Hai, H. T. et al. Whole blood transcriptional profiles and the pathogenesis of tuberculous meningitis. eLife 13, RP92344.

3. Zak, D. E. et al. A blood RNA signature for tuberculosis disease risk: a prospective cohort study. Lancet Lond. Engl. 387, 2312–2322 (2016).

4. Roe, J. et al. Blood Transcriptomic Stratification of Short-term Risk in Contacts of Tuberculosis. Clin. Infect. Dis. 70, 731–737 (2020).

5. Gupta-Wright, A. et al. Evaluation of the Xpert MTB Host Response assay for the triage of patients with presumed pulmonary tuberculosis: a prospective diagnostic accuracy study in Viet Nam, India, the Philippines, Uganda, and South Africa. Lancet Glob. Health 12, e226–e234 (2024).

6. Sweeney, T. E., Braviak, L., Tato, C. M. & Khatri, P. Genome-wide expression for diagnosis of pulmonary tuberculosis: a multicohort analysis. Lancet Respir. Med. 4, 213–224 (2016).

7. Warsinske, H. C. et al. Assessment of Validity of a Blood-Based 3-Gene Signature Score for Progression and Diagnosis of Tuberculosis, Disease Severity, and Treatment Response. JAMA Netw. Open 1, e183779 (2018).

8. Sutherland, J. S. et al. Performance of 2 Finger-Stick Blood Tests to Triage Adults With Symptoms of Pulmonary Tuberculosis: A Prospective Multisite Diagnostic Accuracy Study. Clin. Infect. Dis. Off. Publ. Infect. Dis. Soc. Am. ciaf105 (2025) doi:10.1093/cid/ciaf105.

9. Allwood, B. W. et al. Post-tuberculosis lung health: perspectives from the First International Symposium. Int. J. Tuberc. Lung Dis. Off. J. Int. Union Tuberc. Lung Dis. 24, 820–828 (2020).

10. Maleche-Obimbo, E. et al. Magnitude and factors associated with post-tuberculosis lung disease in low- and middle-income countries: A systematic review and meta-analysis. PLOS Glob. Public Health 2, e0000805 (2022).

11. Ivanova, O., Hoffmann, V. S., Lange, C., Hoelscher, M. & Rachow, A. Post-tuberculosis lung impairment: systematic review and meta-analysis of spirometry data from 1411621 people. Eur. Respir. Rev. 32, 220221 (2023).

12. Zifodya, J. S. et al. Clinical and transcriptomic risk factors for post-tuberculosis lung disease in a cohort of Kenyan adults. Am. J. Respir. Crit. Care Med. aamag063 (2026) doi:10.1093/ajrccm/aamag063.

13. Nathavitharana, R. R. et al. Assessing Infectiousness and the Impact of Effective Treatment to Guide Isolation Recommendations for People With Pulmonary Tuberculosis. J. Infect. Dis. 231, 10–22 (2025).

14. Fennelly, K. P. et al. Cough-generated aerosols of Mycobacterium tuberculosis: a new method to study infectiousness. Am. J. Respir. Crit. Care Med. 169, 604–609 (2004).

15. Nduba, V. et al. Mycobacterium tuberculosis cough aerosol culture status associates with host characteristics and inflammatory profiles. Nat. Commun. 15, 7604 (2024).

16. Zifodya JS et al. Clinical and transcriptomic risk factors for post-tuberculosis lung disease (PTLD) in a cohort of Kenyan adults. AJRCCM In Press,.

17. Fennelly, K. P. et al. Variability of infectious aerosols produced during coughing by patients with pulmonary tuberculosis. Am. J. Respir. Crit. Care Med. 186, 450–457 (2012).

18. van Gemert, F. et al. Prevalence of chronic obstructive pulmonary disease and associated risk factors in Uganda (FRESH AIR Uganda): a prospective cross-sectional observational study. Lancet Glob. Health 3, e44–51 (2015).

19. Jones, P. W. Health status measurement in chronic obstructive pulmonary disease. Thorax 56, 880–887 (2001).

20. Graham, B. L. et al. Standardization of Spirometry 2019 Update. An Official American Thoracic Society and European Respiratory Society Technical Statement. Am. J. Respir. Crit. Care Med. 200, e70–e88 (2019).

21. Quanjer, P. H. et al. Multi-ethnic reference values for spirometry for the 3-95-yr age range: the global lung function 2012 equations. Eur. Respir. J. 40, 1324–1343 (2012).

22. Stanojevic, S. et al. ERS/ATS technical standard on interpretive strategies for routine lung function tests. Eur. Respir. J. 60, 2101499 (2022).

23. Muwanga, V. M. et al. Blood transcriptomic signatures for symptomatic tuberculosis in an African multicohort study. Eur. Respir. J. 64, 2400153 (2024).

24. Mendelsohn, S. C. et al. Transcriptomic Signatures of Progression to Tuberculosis Disease Among Close Contacts in Brazil. J. Infect. Dis. 230, e1355–e1365 (2024).

25. Auld SC, Barczak AK, Bishai W, Coussens AK, Dewi IM, Mitini-Nkhoma SC, Muefong C, Naidoo T, Pooran A, Stek C. Pathogenesis of Post-Tuberculosis Lung Disease: Defining Knowledge Gaps and Research Priorities at the Second International Post-Tuberculosis Symposium. American Journal of Respiratory and Critical Care Medicine 2024; 210:979–993.

26. Tretina, K., Park, E.-S., Maminska, A. & MacMicking, J. D. Interferon-induced guanylate-binding proteins: Guardians of host defense in health and disease. J. Exp. Med. 216, 482–500 (2019).

27. Kirkby, M., Enosi Tuipulotu, D., Feng, S., Lo Pilato, J. & Man, S. M. Guanylate-binding proteins: mechanisms of pattern recognition and antimicrobial functions. Trends Biochem. Sci. 48, 883–893 (2023).

28. Lutz, S. M. et al. A genome-wide association study identifies risk loci for spirometric measures among smokers of European and African ancestry. BMC Genet. 16, 138 (2015).

29. Cho, M. H. et al. Risk loci for chronic obstructive pulmonary disease: a genome-wide association study and meta-analysis. Lancet Respir. Med. 2, 214–225 (2014).

30. Busch, R. et al. Genetic Association and Risk Scores in a Chronic Obstructive Pulmonary Disease Meta-analysis of 16,707 Subjects. Am. J. Respir. Cell Mol. Biol. 57, 35– 46 (2017).

31. Shen, W. et al. RNA-binding protein AZGP1 inhibits epithelial cell proliferation by regulating the genes of alternative splicing in COPD. Gene 927, 148736 (2024).

32. Kim, G.-D., Lim, E. Y. & Shin, H. S. Macrophage Polarization and Functions in Pathogenesis of Chronic Obstructive Pulmonary Disease. Int. J. Mol. Sci. 25, 5631 (2024).

33. Shaykhiev, R. et al. Smoking-dependent reprogramming of alveolar macrophage polarization: implication for pathogenesis of chronic obstructive pulmonary disease. J. Immunol. Baltim. Md 1950 183, 2867–2883 (2009).

34. Folkesson, E. et al. Improved detection of extrapulmonary and paucibacillary pulmonary tuberculosis by Xpert MTB Host Response in a TB low endemic, high resource setting. J. Infect. Dis. jiaf110 (2025) doi:10.1093/infdis/jiaf110.

35. Olbrich, L. et al. Diagnostic accuracy of a three-gene Mycobacterium tuberculosis host response cartridge using fingerstick blood for childhood tuberculosis: a multicentre prospective study in low-income and middle-income countries. Lancet Infect. Dis. 24, 140–149 (2024).

36. Mendelsohn, S. C. et al. Prospective multicentre head-to-head validation of host blood transcriptomic biomarkers for pulmonary tuberculosis by real-time PCR. Commun. Med. 2, 26 (2022).

37. Moreira, F. M. F. et al. Blood-based host biomarker diagnostics in active case finding for pulmonary tuberculosis: A diagnostic case-control study. EClinicalMedicine 33, 100776 (2021).

38. Li, M. et al. Evaluation of the Cepheid 3-gene host response blood test for tuberculosis diagnosis and treatment response monitoring in a primary-level clinic in rural China. J. Clin. Microbiol. 61, e0091123 (2023).

39. Krapp, C. et al. Guanylate Binding Protein (GBP) 5 Is an Interferon-Inducible Inhibitor of HIV-1 Infectivity. Cell Host Microbe 19, 504–514 (2016).

