## supplemental data for "A whole-blood transcriptional signature associated with obstructive post-tuberculosis lung disease"

**Supplemental Materials**

| **Supplemental Table 1. Multivariate Analysis of Modified MTB Host Response Score Association with Pulmonary TB** | | |
| --- | --- | --- |
| Variable | Estimate | p-value |
| GeneXpert cycle threshold | 0.020 | 0.004 |
| MUAC*, (cm) | 0.051 | 7.85e-06 |
| Cavitary CXR | -0.185 | 0.028 |
| Number of affected lung fields | -0.068 | 0.052 |

Presented as median (interquartile range) and number (%), * MUAC=mid upper arm circumference

| **Supplemental Table 2. Cytokine Association Analysis with Modified MTB Host Response Score** | | | |
| --- | --- | --- | --- |
|  |  | estimate | p - value |
| Plasma inflammatory markers | | | |
| CRP* (mg/L, median (IQR)) | 69.1 (33.5 - 105) | -0.004 | 1.05e-12 |
| TNF (median (IQR)) | 16.5 (6.82 - 27.5) | -0.0002 | 0.750 |
| IL6 (152) (median (IQR)) | 22.3 (9.88 - 46.2) | -0.001 | 0.182 |
| Sputum cytokines | | | |
| IL6 (n = 147) (median (IQR)) | 10.8 (2.64 - 52.1) | -0.0004 | 0.109 |
| CXCL8 (n = 147) (median (IQR)) | 3,670 (808 - 7,517) | -2.351e-06 | 0.792 |
| IL1B (n = 146) (median (IQR)) | 699 (191 - 1687) | 1.836e-05 | 0.495 |

Presented as median, (interquartile range) and number (%), *CRP C-reactive protein; TNF=tumor necrosis factor; IL6=interleukin 6; CXCL8= Chemokine (C-X-C motif) ligand 8; IL1B=Interleukin 1B.

| **Supplemental Table 3. Modified MTB Host Response Score and Cough-Aerosol Culture Analysis** | | | | |
| --- | --- | --- | --- | --- |
|  | CASS positive (67)  n (%) or median (IQR) | CASS negative (139)  n (%) or median (IQR) | OR | p value * |
| Age (years) | 31 (25 - 39.5) | 33 (26 - 41) | 0.974 | 0.060 |
| Women | 12 (17.9%) | 34 (24.5%) | 0.674 | 0.292 |
| PLWH* | 9 (17%) | 14 (14.4%) | 1.213 | 0.679 |
| DM* | 8 (11.9%) | 22 (15.9%) | 0.715 | 0.449 |
| HgA1c (mmol/mol) | 5.89 (5.53 - 6.23) | 5.80 (5.48 - 6.17) | 1.022 | 0.817 |
| MUAC* (cm) | 23.4 (21.8 - 25.5) | 22.6 (21.2 - 24.8) | 1.062 | 0.246 |
| BMI* (kg/m^2^) | 18.7 (17.4 - 21.2) | 18.9 (17.7 - 20.9) | 1.008 | 0.874 |
| AFB* Smear positive | 62 (92.5%) | 122 (87.8%) | 1.728 | 0.304 |
| GeneXpert CT* | 17.2 (15.9 - 18) | 18.4 (17.1 - 23.9) | 0.777 | 3.27e-05 |
| Cavitary disease | 53 (79.1%) | 84 (60.4%) | 2.479 | 0.009 |
| CRP (mg/L) | 82.6 (42.8 - 113) | 54.3 (26.3 - 92.4) | 1.006 | 0.032 |
| CXR severity (quadrants) | 2 (1 - 3) | 2 (1 - 3) | 1.082 | 0.572 |
| mHR Score | -0.103 (-0.412  - 0.162) | -0.006 (-0.332 - 0.586) | 0.486 | 0.009 |
| Change in mHR after 6 months | 1.3 (0.997 - 1.7) | 1.37 (0.813 - 1.79) | 1.146 | 0.712 |

Presented as median (interquartile range) and number (%), *AFB=acid-fast bacilli; BMI=body mass index; CT=cycle threshold; CXR=chest X-ray; MUAC=mid upper arm circumference; DM=Diabetes Mellitus; PLWH=persons living with HIV; mHR=modified MTB Host Response score.

| **Supplemental Table 4. Multivariate analysis of Modified MTB Host Response Score Cough-Aerosol Culture Positivity** | | |
| --- | --- | --- |
|  | OR | p-value |
| GeneXpert Cycle threshold | 0.792 | 8.93e-05 |
| mHR score | 0.649 | 0.198 |
| Cavitary disease | 1.451 | 0.366 |
| Age (years) | 0.983 | 0.322 |

| **Supplemental Table 5. Modified MTB Host Response Score Association with Overall PTLD and Restrictive PTLD at 6 and 12 Months.** | | | | | |
| --- | --- | --- | --- | --- | --- |
| A. 6 months |  |  |  |  |  |
|  | Normal PFTs n = 110 | PTLD n = 106 | p-value | Restriction n = 69 | p-value |
| Age(years) | 32.5 (25 - 43) | 33 (26 - 43) | 0.638 | 33 (26 - 44) | 0.922 |
| Women | 25 (22.7%) | 21 (19.8%) | 0.601 | 14 (20.3%) | 0.701 |
| PLWH* | 10 (12.2%) | 12 (14.3%) | 0.692 | 5 (9.09%) | 0.570 |
| DM* | 12 (10.9%) | 12 (11.3%) | 0.923 | 7 (10.1%) | 0.872 |
| HgA1c(mmol/mol) | 5.7 (5.45 - 6.04) | 5.68 (5.3 - 6.07) | 0.944 | 5.61 (5.29 - 6.02) | 0.924 |
| MUAC(cm) | 23.5 (22 - 25.5) | 22 (20.5 - 24) | 5.97e-04 | 21.9 (20 - 23) | 1.21e-04 |
| BMI(kg/m^2^) | 19.8 (18.2 - 22.1) | 18.2 (16.8 - 20.2) | 1.44e-04 | 18 (16.6 - 19) | 5.94e-06 |
| AFB Smear positive | 90 (81.8%) | 97 (91.5%) | 0.444 | 64 (92.8%) | 0.444 |
| GeneXpert CT | 18.8 (17.2 - 24) | 18.4 (17.3 - 21.6) | 0.109 | 18.0 (17.3 - 21.2) | 0.126 |
| Cavitary disease | 17 (15.5%) | 20 (18.9%) | 0.506 | 14 (20.3%) | 0.407 |
| CRP(mg/L) | 49.8 (15.1 - 100) | 58.3 (24.3 - 106) | 0.516 | 66.8 (40.9 - 109) | 0.101 |
| CXR severity (quadrants) | 2 (1 - 2) | 2 (1 - 3) | 1.07e-05 | 2 (1 - 3) | 6.68e-05 |
| mHR Score | 0.037 (-0.334 - 0.581) | 0.063 (-0.280 - 0.479) | 0.685 | -0.173 (-0.550 - 0.139) (40) | 0.057 |
| Change in mHR after 6 months | 1.33 (0.868 - 1.62) | 1.23 (0.948 - 1.46) | 0.435 | 1.13 (0.648 - 1.42) | 0.164 |
| B. 12 months |  |  |  |  |  |
|  | Normal PFTs n = 105 | PTLD n = 105 | p-value | Restriction n = 61 | p-value |
| Age(years) | 31 (25 - 44) | 33 (25 - 43) | 0.944 | 32 (25 - 40) | 0.383 |
| Women | 26 (24.8%) | 22 (21.0%) | 0.511 | 14 (23.0%) | 0.793 |
| PLWH* | 10 (12.%) | 9 (10.5%) | 0.745 | 5 (10%) | 0.718 |
| DM* | 10 (9.52%) | 6 (5.71%) | 0.303 | 6 (9.84%) | 0.948 |
| HgA1c(mmol/mol) | 5.56 (5.3 - 5.98) | 5.63 (5.22 - 5.94) | 0.287 | 5.57 (5.19 - 5.9) | 0.432 |
| MUAC(cm) | 23.5 (21.9 - 25.4) | 22.5 (20.9 - 24.2) | 0.017 | 22.4 (21 - 23.8) | 0.054 |
| BMI(kg/m^2^) | 19.5 (17.8 - 21.3) | 18.5 (17.1 - 20.6) | 0.062 | 18.4 (16.9 - 20.6) | 0.153 |
| AFB Smear positive | 89 (84.8%) | 95 (90.5%) | 0.444 | 55 (90.2%) | 0.444 |
| GeneXpert CT | 19.4 (16.9 - 24.2) | 18 (16.6 - 20.7) | 0.072 | 17.9 (17.2 - 21.1) | 0.124 |
| Cavitary disease | 20 (19.0%) | 25 (23.8%) | 0.401 | 13 (21.3%) | 0.725 |
| CRP(mg/L) | 53.2 (15.1 - 91.8) | 66.8 (27.1 - 104) | 0.127 | 73.5 (36.5 - 111) | 0.024 |
| CXR severity (quadrants) | 1 (1 - 2) | 2 (2 - 3) | 1.28e-07 | 2 (2 - 3) | 2.46e-05 |
| mHR Score | -0.174 (-0.498 - 0.541) (61) | 0.033 (-0.292 - 0.238) | 0.604 | -0.124 (-0.405 - 0.160) (39) | 0.548 |
| Change in mHR after 6 months | 1.33 (0.868 - 1.62) | 1.23 (0.948 – 1.46) | 0.671 | 1.22 (0.745 - 1.41) | 0.420 |

Presented as median (interquartile range) and number (%), *AFB=acid-fast bacilli; BMI=body mass index; CT=cycle threshold; CXR=chest X-ray; MUAC=mid upper arm circumference; DM=Diabetes Mellitus; PLWH=persons living with HIV; mHR=modified MTB Host Response score.

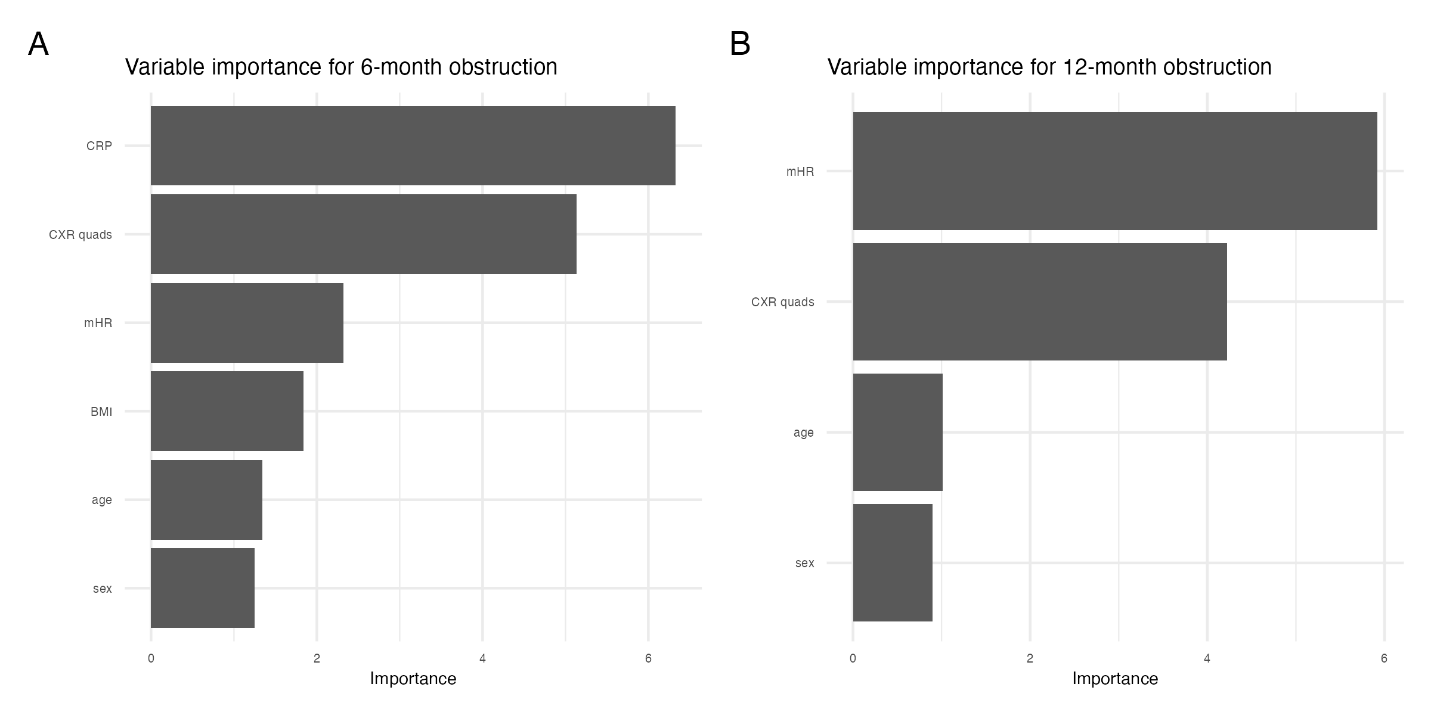

**Supplemental Figure 1:** Variable importance plots using variables associated with obstructive PTLD with p ≤ 0.1 at (A) 6 and (B) 12 months. Age and sex were also included due to biological plausibility. Variables with importance ≥ 1 were used in the final obstructive PTLD prediction analysis.
